# International Comparisons of Harmonized Laboratory Value Trajectories to Predict Severe COVID-19: Leveraging the 4CE Collaborative Across 342 Hospitals and 6 Countries: A Retrospective Cohort Study

**DOI:** 10.1101/2020.12.16.20247684

**Authors:** Griffin M Weber, Chuan Hong, Nathan P Palmer, Paul Avillach, Shawn N Murphy, Alba Gutiérrez-Sacristán, Zongqi Xia, Arnaud Serret-Larmande, Antoine Neuraz, Gilbert S. Omenn, Shyam Visweswaran, Jeffrey G Klann, Andrew M South, Ne Hooi Will Loh, Mario Cannataro, Brett K Beaulieu-Jones, Riccardo Bellazzi, Giuseppe Agapito, Mario Alessiani, Bruce J Aronow, Douglas S Bell, Antonio Bellasi, Vincent Benoit, Michele Beraghi, Martin Boeker, John Booth, Silvano Bosari, Florence T Bourgeois, Nicholas W Brown, Mauro Bucalo, Luca Chiovato, Lorenzo Chiudinelli, Arianna Dagliati, Batsal Devkota, Scott L DuVall, Robert W Follett, Thomas Ganslandt, Noelia García Barrio, Tobias Gradinger, Romain Griffier, David A Hanauer, John H Holmes, Petar Horki, Kenneth M Huling, Richard W Issitt, Vianney Jouhet, Mark S Keller, Detlef Kraska, Molei Liu, Yuan Luo, Kristine E Lynch, Alberto Malovini, Kenneth D Mandl, Chengsheng Mao, Anupama Maram, Michael E Matheny, Thomas Maulhardt, Maria Mazzitelli, Marianna Milano, Jason H Moore, Jeffrey S Morris, Michele Morris, Danielle L Mowery, Thomas P Naughton, Kee Yuan Ngiam, James B Norman, Lav P Patel, Miguel Pedrera Jimenez, Rachel B Ramoni, Emily R Schriver, Luigia Scudeller, Neil J Sebire, Pablo Serrano Balazote, Anastasia Spiridou, Amelia LM Tan, Byorn WL Tan, Valentina Tibollo, Carlo Torti, Enrico M Trecarichi, Michele Vitacca, Alberto Zambelli, Chiara Zucco, The Consortium for Clinical Characterization of COVID-19 by EHR (4CE), Isaac S Kohane, Tianxi Cai, Gabriel A Brat

## Abstract

**Objectives:** To perform an international comparison of the trajectory of laboratory values among hospitalized patients with COVID-19 who develop severe disease and identify optimal timing of laboratory value collection to predict severity across hospitals and regions.

**Design:** Retrospective cohort study.

**Setting:** The Consortium for Clinical Characterization of COVID-19 by EHR (4CE), an international multi-site data-sharing collaborative of 342 hospitals in the US and in Europe.

**Participants:** Patients hospitalized with COVID-19, admitted before or after PCR-confirmed result for SARS-CoV-2.

**Primary and secondary outcome measures:** Patients were categorized as “ever-severe” or “never-severe” using the validated 4CE severity criteria. Eighteen laboratory tests associated with poor COVID-19-related outcomes were evaluated for predictive accuracy by area under the curve (AUC), compared between the severity categories. Subgroup analysis was performed to validate a subset of laboratory values as predictive of severity against a published algorithm. A subset of laboratory values (CRP, albumin, LDH, neutrophil count, D-dimer, and procalcitonin) was compared between North American and European sites for severity prediction.

**Results:** Of 36,447 patients with COVID-19, 19,953 (43.7%) were categorized as ever-severe. Most patients (78.7%) were 50 years of age or older and male (60.5%). Longitudinal trajectories of CRP, albumin, LDH, neutrophil count, D-dimer, and procalcitonin showed association with disease severity. Significant differences of laboratory values at admission were found between the two groups. With the exception of D-dimer, predictive discrimination of laboratory values did not improve after admission. Sub-group analysis using age, D-dimer, CRP, and lymphocyte count as predictive of severity at admission showed similar discrimination to a published algorithm (AUC=0.88 and 0.91, respectively). Both models deteriorated in predictive accuracy as the disease progressed. On average, no difference in severity prediction was found between North American and European sites.

**Conclusions:** Laboratory test values at admission can be used to predict severity in patients with COVID-19. Prediction models show consistency across international sites highlighting the potential generalizability of these models.

## INTRODUCTION

Severe acute respiratory syndrome coronavirus 2 (SARS-CoV-2) has caused millions of cases of coronavirus disease 2019 (COVID-19) in nearly every country. While most patients with COVID-19 have a mild form of viral pneumonia, an appreciable subgroup develops rapid onset of severe disease. Several large national studies have demonstrated that a variable and potentially significant proportion (ranging from 5-70%) [1–3] of hospitalized patients with COVID-19 develop cardiorespiratory failure, require mechanical ventilation and hemodynamic support, and may ultimately die. The early identification of patients at high risk for severe disease and worse outcomes can improve triage and resource allocation, particularly when numbers of COVID-19 cases overwhelm health systems. [4]

Numerous studies have reported models using clinical data, including laboratory values, to predict patients at high risk of severe disease. [2] However, most models have not been tested across hospital systems and countries to determine generalizability. Few studies have included patients from multi-national cohorts.

We formed the 4CE Consortium [5] as an international research collaborative of nearly 350 hospitals from six countries in order to collect standardized patient-level electronic health record (EHR) data to examine the epidemiology, pathophysiology, management, and healthcare system dynamics of COVID-19. Using the 4CE data, we examined the relationship between pre-selected laboratory values [6] collected during the early phase of hospitalization for COVID-19 and subsequent progression to severe disease during hospitalization across institutions and countries. We compared prediction models using single laboratory values at admission and during hospitalization to a prediction model containing multiple laboratory values. We chose the model developed by Chen and Liu [7] as it included laboratory values we could easily compare. Across all models, we evaluated geographical differences (national and continental) among the severity prediction models.

## METHODS

### Cohort identification

We included all patients hospitalized at participating 4CE sites with an admission date from 7 days before to 14 days after the date of their first reverse transcription polymerase chain reaction (PCR)-confirmed SARS-CoV-2 positive test result. The first admission date within this 21-day time window was considered the index admission date. Throughout this work, “days since admission” refers to this index date.

### Participating sites

Data were available from 36,447 patients from 342 hospitals (affiliated with 46 sites) across six countries: France, Germany, Italy, Singapore, Spain, and the United States. See eTABLE 2 for details about participating sites. Several sites collected data from multiple hospitals. In the United States, 170 medical centers of the US Department of Veterans Affairs were grouped into 18 regional divisions called Veterans Integrated Service Networks.

### Patient and Public Involvement

Patients and the public were not involved in the design, conduct, or reporting, or dissemination plans of the research.

### Severe COVID-19

We categorized the primary patient outcome as “ever-severe” or “never-severe” based on whether the patients, at any time during their hospitalization, progressed to severe disease—even if they later recovered from COVID-19. We defined “ever-severe” COVID-19 using the validated 4CE severity criteria based on the presence of at least one of the following codes in the EHR during the hospitalization: an order for any medication in two broad classes (intravenous anesthetics or cardiac inotropes), a collected partial pressure of arterial oxygen (PaO2) laboratory test (regardless of the result), an International Classification of Disease (ICD)-10 diagnosis code for acute respiratory distress syndrome or ventilator-associated pneumonia, or an ICD-10 procedure code for insertion of endotracheal tube or invasive mechanical ventilation [8]. A more complete description is available in eTABLE 3. The full list of codes is available in [8] and in the consortium’s GitHub [9].

We chose this definition of severity because of the availability of standardized coded data describing medications, laboratory tests, diagnoses, and procedures. In contrast, other measures of disease severity, including requirement for mechanical ventilation or intensive care and death, are often unavailable in the EHR or are inconsistently recorded and are not easily ascertained. As an example, admission to traditional ICU spaces was not an effective measure of ICU admission when surge conditions required non-traditional placement.

### Local data collection and central data aggregation

Following our prior approach [5], each contributing site executed queries on local database systems containing EHR data to generate six tables in comma-separated values (CSV) format, containing aggregate counts and statistics on their respective patient cohorts: DailyCounts, ClinicalCourse, Demographics, Labs, Diagnoses, and Medications (see eTABLE 4 for descriptions of the files and Figure 1). Sites uploaded their six CSV files to a central 4CE Data Upload Tool, which performed several validation steps, which included variable consistency and range and count evaluation. The results of validation were either a report of any errors or the ability to save the files to a private shared folder. Throughout this process, patient-level files remained at each site and were not centrally shared. A key advantage of executing queries locally and sharing only aggregate data was that sites were able to obtain institutional approval more rapidly. Further, this approach facilitated analysis at the site, country, and region by reducing barriers to federated data generation and aggregation at each level.

**FIGURE 1.**
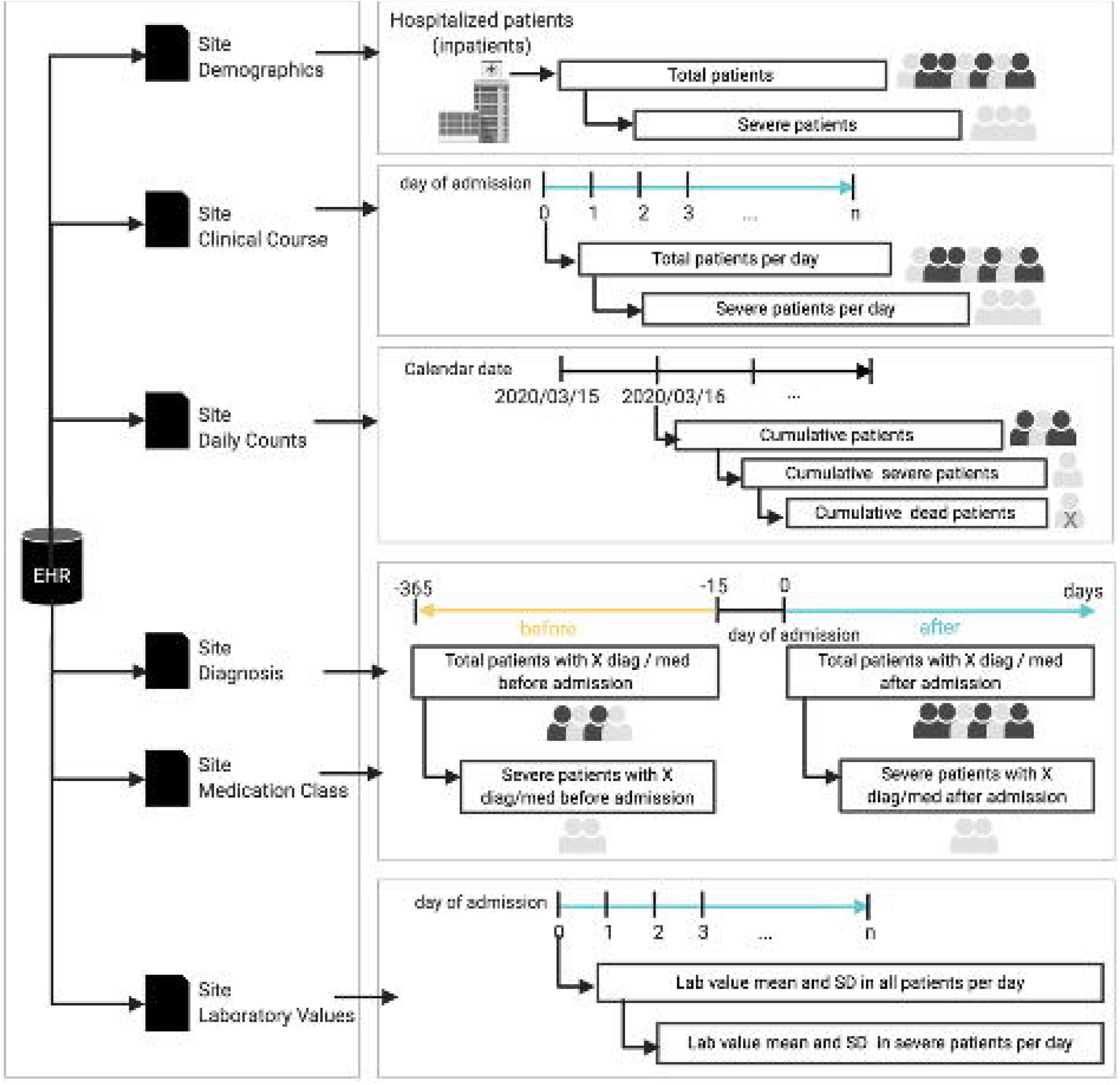
Each site generated six data tables (comma-separated files) containing aggregate counts and statistics on their individual level data: 1) demographic breakdowns, 2) clinical course shown as counts and disposition from index date, 3) daily counts of patients and their disposition, 4) daily diagnosis counts and 5.) daily medication counts 6) daily trajectories of lab tests. These aggregate descriptive files without individual level data were provided to the consortium for extensive quality-assurance steps (see Methods).

Most sites used the open-source i2b2 (Informatics for Integrating Biology and the Bedside) software platform to obtain the data. More than 200 organizations worldwide use i2b2 for purposes that include identifying participants for clinical trials, drug safety monitoring, and clinical and epidemiological research. Those 4CE sites with i2b2 used database scripts to directly query their i2b2 repository, calculate the counts and statistics, and export the data files. The 4CE sites without i2b2 used the Observational Medical Outcomes Partnership (OMOP) Common Data Model or their own clinical data warehouse solutions (e.g., Epic Caboodle) and querying tools to create the required files.

### Selection of laboratory tests and medications

We focused on 18 laboratory tests associated with worse outcomes in patients with COVID-19 based on prior reports [6]. We provided each site with a single standard Logical Objects, Identifiers, Names and Codes (LOINC) identifier for each test, but sites often needed to map tests to additional LOINC or custom codes within their EHR. We addressed barriers that arose during initial efforts to extract these laboratory values by stratifying region-specific laboratory test types to reduce extraction errors and enable standardization. For example, D-dimer was extracted with both fibrinogen-equivalent unit and D-dimer unit measurements. We converted all results to D-dimer units for subsequent analyses.

### Quality control

After file upload, the online validation process checked the file and column names, column orders, data types, code values and ranges, and duplicated records. Then, we ran an R script for the following additional quality control checks: consistency of the total counts of ever-severe and never-severe cases across all datasets within each site, consistency between the 3-digit diagnosis codes and the ICD dictionary, and consistency of the range of laboratory data from each site with data observed from all sites. We contacted a site for site-specific quality control if the site laboratory values were consistently lower or higher than the other sites or otherwise implausible.

### Statistical analysis

We estimated the country-level daily incidence of new patients hospitalized with COVID-19 during the study period from January 23, 2020 to September 29, 2020. Specifically, for each country, we summed the daily incidence of new patients hospitalized with COVID-19 at each site within that country per 100,000 people of the country and multiplied this by an adjustment factor, defined as the ratio between the country’s overall inpatient discharge rate and the overall inpatient discharge rate of all 4CE sites in that country irrespective of COVID-19 status. We then reported the adjusted 7-day average incidence of new COVID-19 hospitalizations per 100,000 of the country population.

We assessed the performance of laboratory values in predicting severity. We summarized the severity risk among different demographic subgroups by country based on the DerSimonian and Laird random effects meta-analysis of the site-specific risk estimates [10]. For each laboratory test, we first compared the mean values over time among the ever-severe group to those in the never-severe group. To account for cross-site variabilities and mitigate confounding bias, we summarized the overall mean trajectory over time via random effects meta-analyses and only included sites that had measurements for patients in both the ever-severe and never-severe groups.

For each laboratory test, we assessed the ability of laboratory results measured at sequential days following the admission date to predict severity risk. We quantified the discriminatory capacity of each laboratory result by estimating the area under the receiver operating characteristic curve (AUC), assuming that the laboratory values within the ever-severe group or the never-severe group at each site followed either a normal distribution or a log-normal distribution. The AUC estimate quantified the probability that the laboratory result of a randomly selected patient in the ever-severe group was sufficiently different from that of a randomly selected patient in the never-severe group. For each laboratory value measured on a given day, we obtained AUC estimates within each site and combined them across sites using meta-analysis (see Supplementary Materials). In addition, we estimated site-level AUCs as the probability that the average laboratory value in an ever-severe group selected randomly from all sites is higher than the average laboratory value in a never-severe group selected randomly from all sites. Site-level AUC assessed the degree to which each laboratory value is different *on average* in the ever-severe group compared to the never-severe group.

Recent literature suggested that combining multiple laboratory measurements may yield better severity predictions. We chose to validate a published composite severity risk score, calculated as a weighted average of several laboratory measurements, in comparison to our individual laboratory predictions. We chose [7] given the extremely high AUC, recent publication, and ease of application in our data set.

## RESULTS

### Characteristics of the Study Population

In this study population of 36,447 patients, the incidence of hospitalization for COVID-19 largely tracked with population dynamics of COVID-19 cases [11] across different countries during the initial pandemic period (FIGURE 2). Both the COVID-19 case rate and the COVID-19 hospitalization rate dropped significantly from the first peak in April 2020. While hospitalization rates and incidence of severe disease remained relatively low for all countries, case rates increased in France, United States, and Spain and Singapore after June 2020. Hospitalization rate and incidence of ever-severe disease in Spain and Singapore increased after August 2020.

**FIGURE 2.**
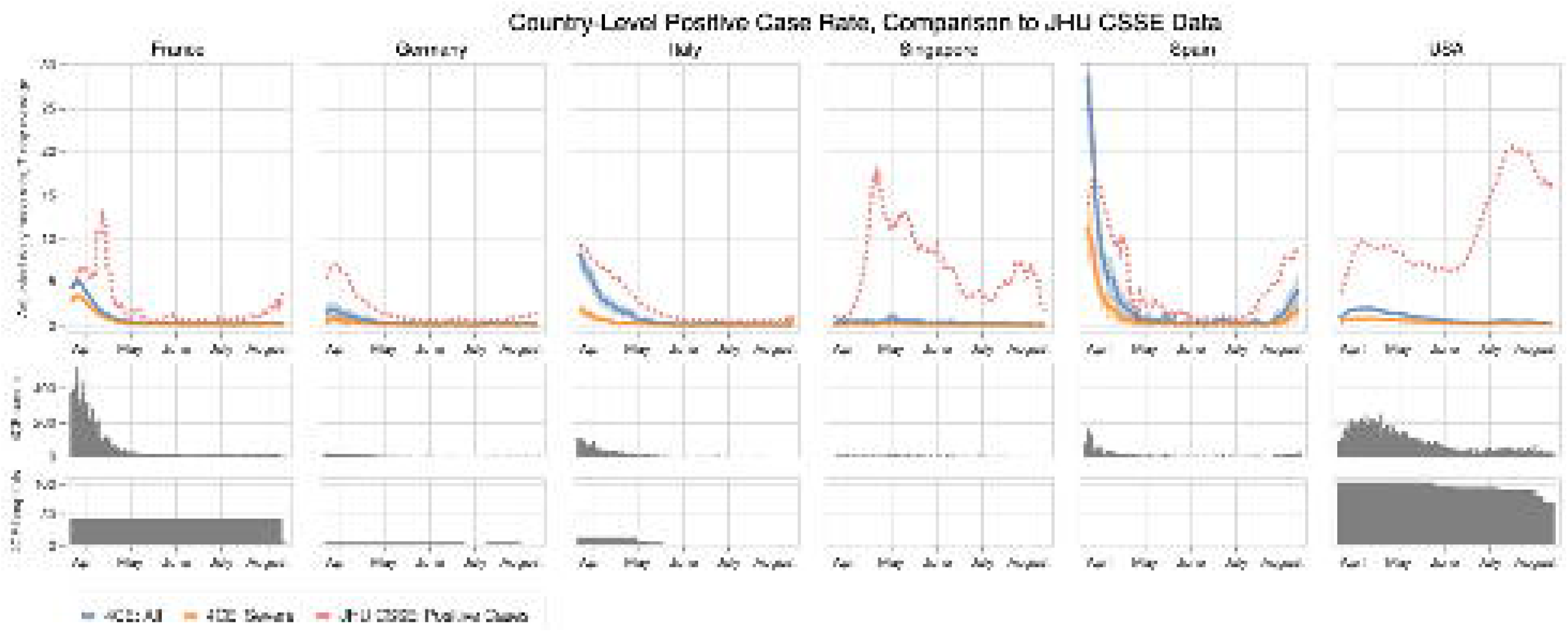
Adjusted 7-day average new hospitalization rate and rate of ever-severe disease per 100,000 people by country based on 4CE contributors along with 95% confidence intervals compared with 7-day average new case rates collected by Johns Hopkins Center for Systems Science and Engineering (JHU CSSE).

Consistent with prior studies [4][12], the study population of patients hospitalized with COVID-19 showed a higher prevalence of men and older populations. See Table 1 for demographic characteristics among ever-severe and never-severe patients and percentages among age group, race/ethnicity, and sex. In our sample, men in the 50-69 age group made up a plurality (25%) of admitted cases, were the most likely to have ever-severe disease. International comparisons were consistent and showed across six countries that older age was associated with increased risk of severe disease (eFIGURE 1). In the United States, where race/ethnicity data were available, minority patients had a higher number of COVID-19 hospitalizations and proportion of severe disease relative to the White reference group.

**TABLE 1.**
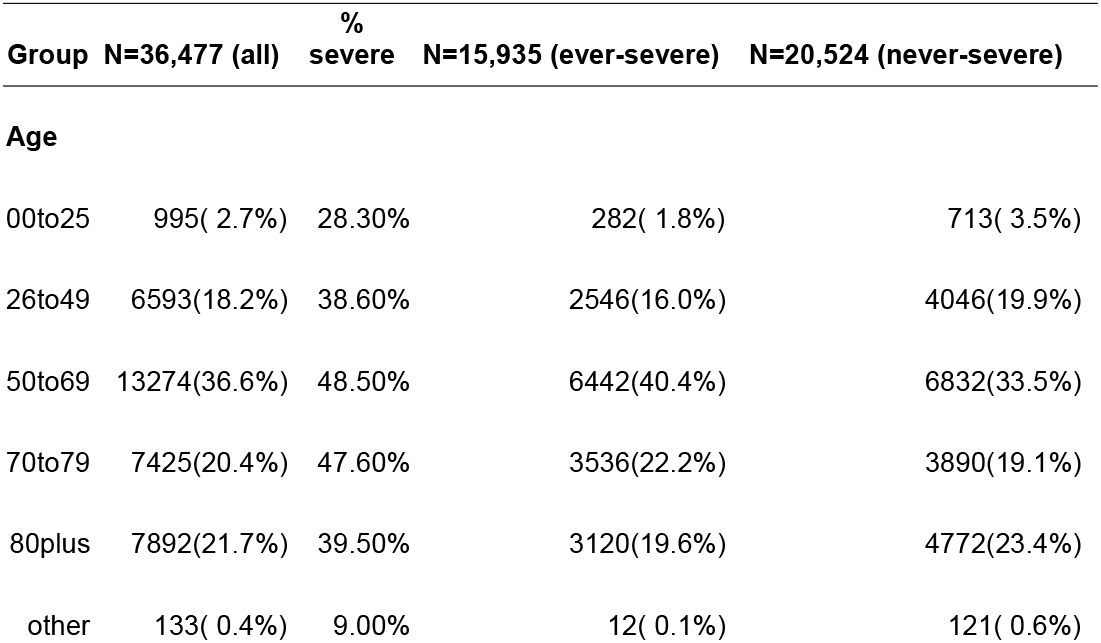
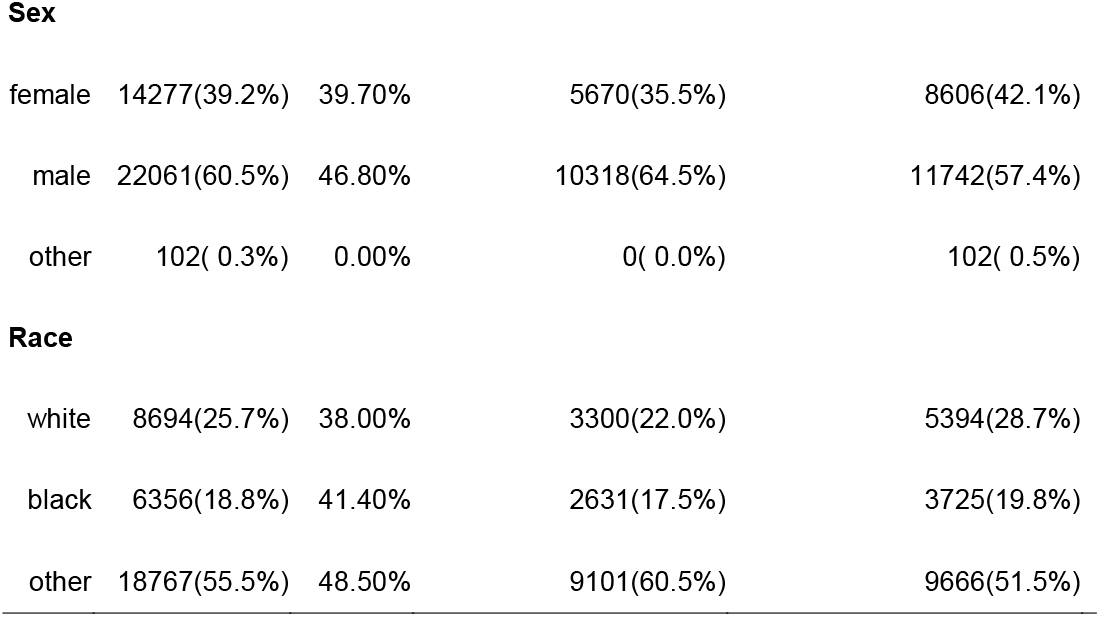
Demographic characteristics among ever-severe and never-severe patients.

## LABORATORY TRAJECTORIES

We evaluated the predictive performance of individual and combined laboratory values available at admission and throughout hospitalization to predict our measure of severe disease. Specifically, we investigated the longitudinal trajectories, AUCs, and thresholds of eighteen laboratory values that were associated with severe disease in prior studies. Six of the labs were most robustly replicated in our analysis: C-reactive protein (CRP), albumin, lactate dehydrogenase (LDH), neutrophil count, D-dimer, and procalcitonin, as shown in FIGURE 3. Trajectories for the remaining laboratory values are shown in the supplemental data (eFIGURE 2).

**FIGURE 3.**
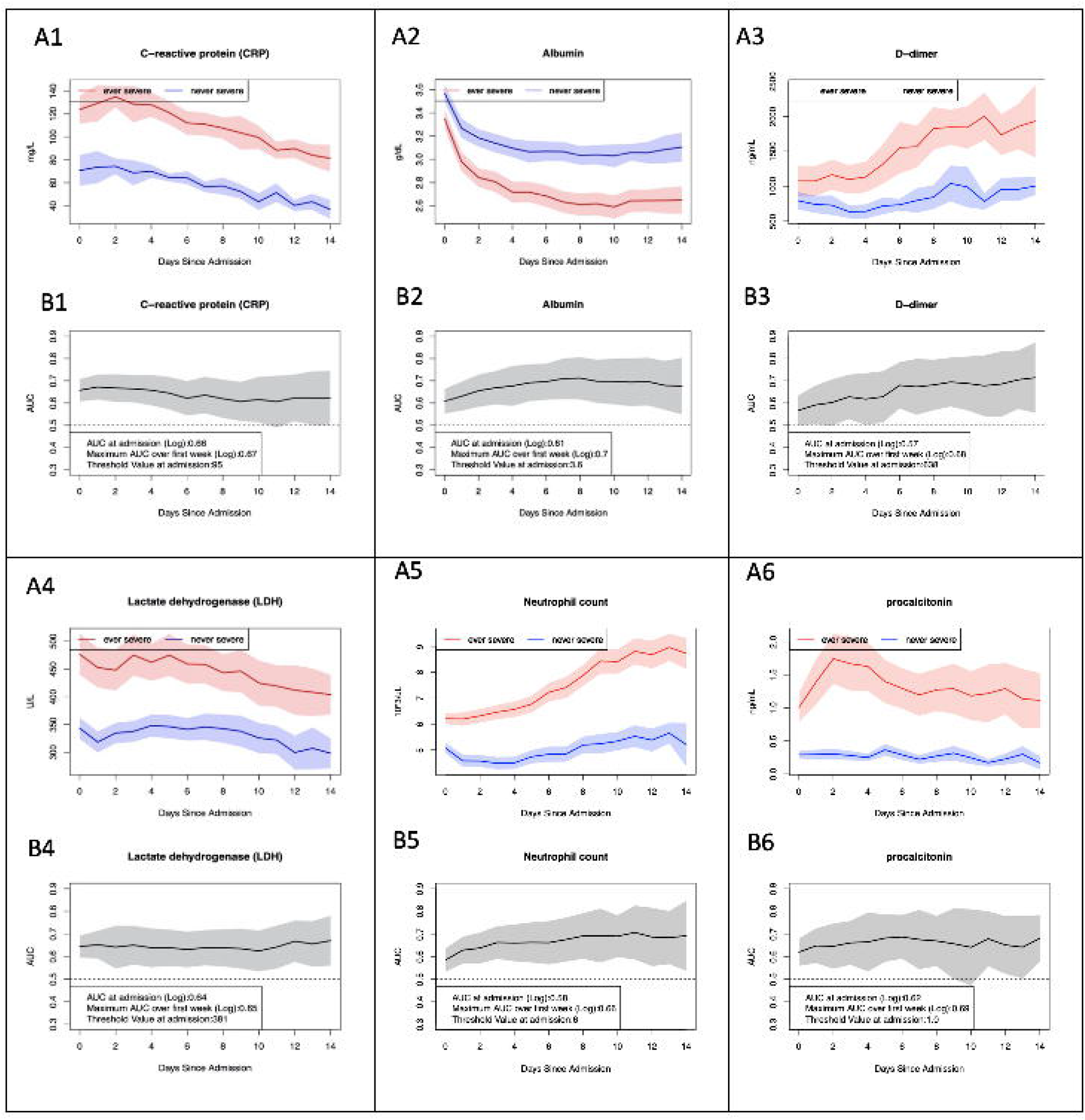
A) Pooled laboratory values in ever-severe and never-severe patients for six selected laboratory tests, and B) patient-level AUC at each day after admission for those labs. Inset shows AUC of laboratory value at admission and in-hospital to individually predict ever-severe as well as optimized thresholds.

When examining the distribution of between-site variation relative to average within-site variation stratified by country across all laboratory tests over 7-day windows, we found that the relative variation was <1, indicating that the site-level variations were much larger than the country-level variations (see eFIGURE 3). Using country-level data to evaluate prediction scores would incorrectly assume consistency of presentation of patients at each site within a country. As a result, we focused on site-level analysis of laboratory values thereafter.

### Predicting Severity from Individual Laboratory Values at Admission

We pooled admission laboratory measures in ever-severe and never-severe patients across all sites (eTABLE 5). After log transformation, differences in admission laboratory values were significant for all laboratory tests (FIGURE 3). For most laboratory studies, differences in initial laboratory values widened between ever-severe and never-severe cases during the first few days after admission. For example, average CRP at admission across all sites was 124 mg/dL (95% CI of 111-135) for ever-severe patients and 71 mg/dL (95% CI of 57-84) for never-severe patients. Initial CRP values predicted severe disease with an AUC of 0.66 (95% CI of 0.61-0.71). CRP was the best laboratory test for distinguishing severe patients at admission in our study, with a threshold value of 91 mg/dL. Additional laboratory studies with potential ability to identify severe disease at admission were D-dimer (AUC = 0.57, 95% CI = 0.50-0.63), albumin (AUC = 0.61, 95% CI = 0.55-0.66), LDH (AUC = 0.64, 95% CI = 0.59-0.69), and procalcitonin (AUC = 0.62, 95% CI = 0.56-0.68).

### Predicting Severity using Individual Laboratory Values after Admission

Given the potential value of laboratory values to better predict patients who transition to severe disease, we next examined the predictive performance of the laboratory values during the hospitalization (FIGURE 3, eFIGURE 2). Laboratory values exhibited distinct trajectories over time. For instance, CRP rapidly fell after an initial peak shortly after admission. In contrast, neutrophil count rose gradually during the hospitalization and ultimately plateaued. Confidence intervals widened over time for all laboratory values due to decreasing sample size with prolonged hospitalization.

We calculated AUC (and 95% CI) of six laboratory tests that distinguished ever-severe and never-severe patients over 14 days. Albumin and CRP were the best predictors of ever-severe disease. Compared to laboratory values on admission, prediction models based on CRP values over the 7 days after admission only marginally improved prediction. Maximum AUC for CRP and albumin was 0.66 and 0.68, respectively. Procalcitonin, neutrophil count, and LDH also had an AUC greater than 0.6. Thus, the utility of most laboratory tests in predicting severe disease from measurements subsequent to admission was minimal. One notable exception was D-dimer. Its performance in predicting severe disease improved from AUC=0.56 (95% CI of 0.50 - 0.63) at admission to AUC=0.68 (95% CI of 0.58 - 0.77) during the later stage of the hospitalization, suggesting that initial elevations of the disease may be less valuable for predicting ever-severe disease.

### Validating Existing Risk Scores

Next, we performed a subgroup analysis to assess whether a combination of laboratory tests would improve prediction of severe disease over each individual laboratory test. Specifically, we chose to validate a published algorithm [7] that included age, D-dimer, CRP, and lymphocyte count. For the three laboratory tests, we used site-level average values. FIGURE 4 shows the performance of the algorithm to identify severe cases over the first 14 days of hospitalization. For the subset of 11 sites where we had all necessary admission laboratory data, we found that the algorithm had an AUC of 0.91 at day 0, which was similar to the published AUC of 0.881. Similar to models based on individual laboratory tests, prediction performance improved marginally in the first couple days after admission and then gradually deteriorated over time. Such changes in performance underlined the need for prediction systems that explicitly address early versus later periods.

**FIGURE 4.**
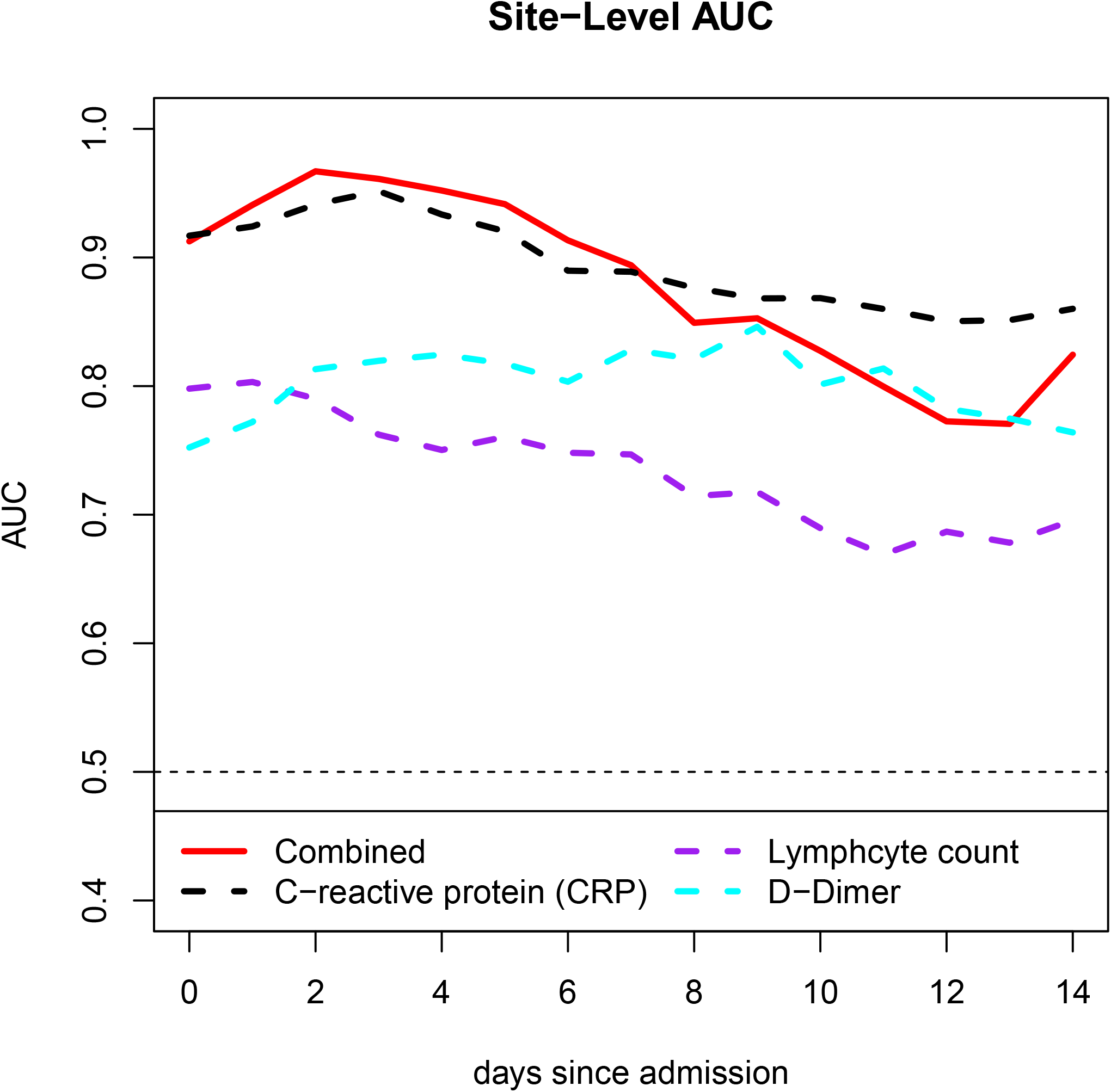
Site level AUC of the risk score compared to the individual laboratory tests.

### Evaluating Geographical Differences in Severity Prediction

Given the unique multi-national nature of the 4CE consortium, we compared performance of the best performing laboratory tests (CRP, albumin, LDH, neutrophil count, D-dimer, and procalcitonin) over time between continents, after grouping North American and European sites (FIGURE 5). Overall, the predictive performance was similar between the sites from the two continents at admission (e.g., for CRP at North American sites: AUC=0.63, 95% CI 0.56 - 0.72; European sites: AUC=0.67, 95% CI 0.63 - 0.70) and over time (e.g., for CRP at North American sites: AUC=0.65, 95% CI 0.56 - 0.74; European sites: AUC=0.68, 95% CI 0.65 - 0.72). We observed a similar pattern for other laboratory tests (eFIGURE4). While variation in laboratory results was notably greater in North America than Europe at all time points, the average results between these two continents were surprisingly similar. These results indicate that it is feasible to use national datasets for severity prediction.

**FIGURE 5.**
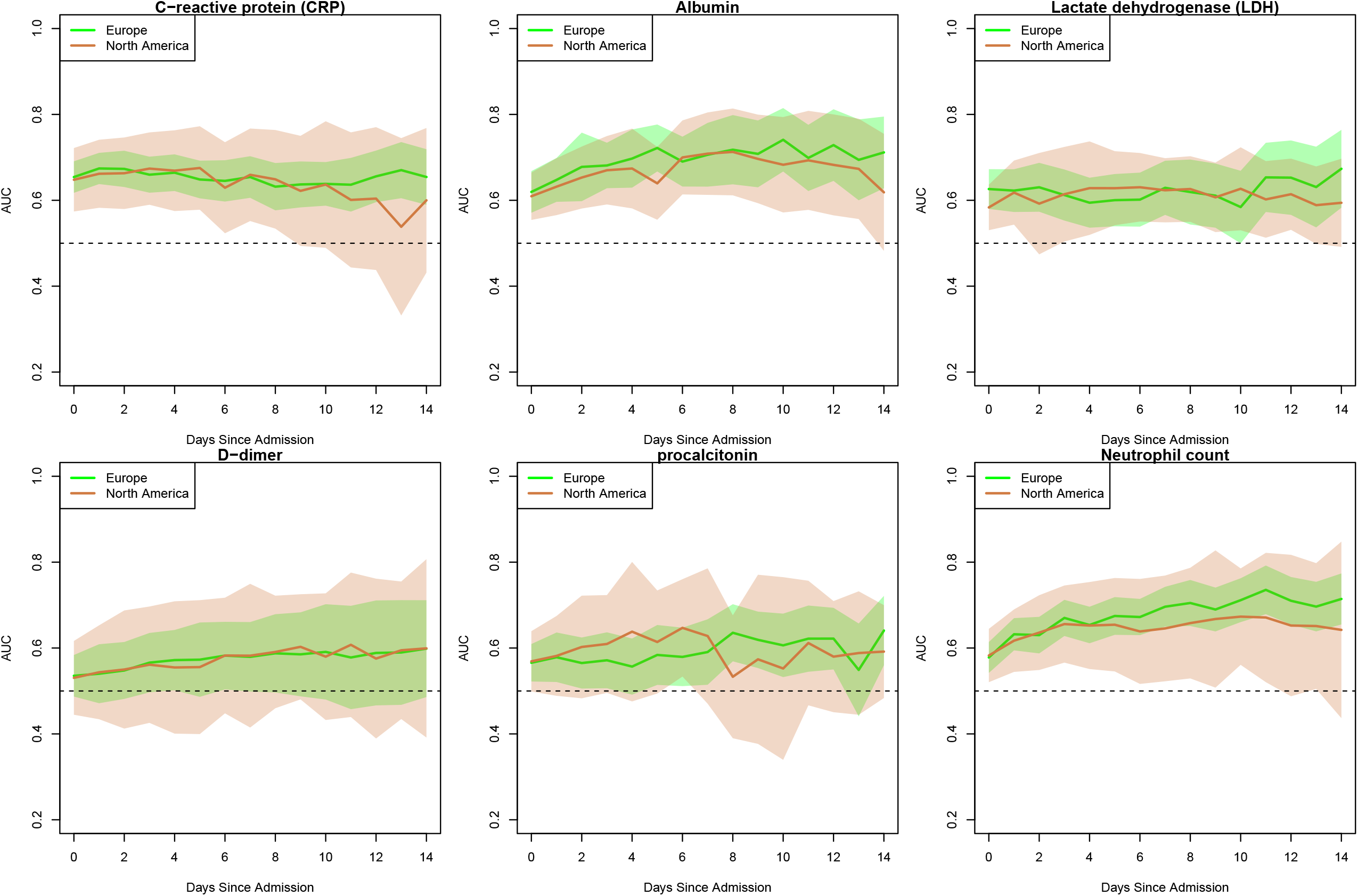
Patient level AUC of six selected laboratory tests stratified by regions.

## DISCUSSION

### Statement of Principal findings

This study builds upon the growing literature of COVID-19 severity prediction to report key findings pertaining to the optimal timing of inpatient laboratory results to predict a severe course of disease. We additionally used an international cohort to validate the generalizability of these models. First, we identified severe patients using a previously validated algorithm within the 4CE consortium. For patients hospitalized for COVID-19, we identified six laboratory tests with the highest predictive power to identify severe disease: CRP, LDH, procalcitonin, D-dimer, neutrophil count, and albumin. These laboratory tests are a subset of a list of tests previously associated with worse outcomes in patients with COVID-19.[6] Second, with the exception of D-dimer, laboratory values at the time of admission showed comparable predictive performance to values measured 7 days after admission. These findings suggest the predictive performance of the identified individual laboratory tests is not significantly improved after admission. Third, we reported laboratory value thresholds for these multiple laboratory tests to predict transitions to severe disease which should be validated with individual-level patient data in future work. Finally, we report important comparisons of laboratory studies between North American and European sites.

### Strengths and weaknesses

The nature and construction of the 4CE consortium offers a number of key strengths and requires acknowledgement of important limitations. The consortium approach enabled the pooling of laboratory values across 342 hospitals with diverse healthcare practices. This showed that site-level (within-site and between-site) differences were greater than country-level differences. Interestingly, the severity predictive performance (AUC) of each laboratory study is remarkably consistent between North America and Europe. Despite the differences in the composition of ever-severe and never-severe patients across sites and countries, the directionality of laboratory values and the threshold for ever-severe disease shared many similarities. The design of the consortium and these analyses highlight that these findings are unlikely to be site-specific or the result of health care system dynamics; the predictive nature of identified lab tests is more likely to generalize. Additionally, given the diversity of sites, the findings are unlikely to be biased by a majority population demographic.

The federated nature of analyses presented several additional limitations the consortium acknowledged and took measures to mitigate. First, EHR data have intrinsic noise, variable levels and causes of missing data, and policy effects on available documentation that will result in differences between sites. By leveraging a federated system of common EHR data elements and capturing site-level heterogeneity to identify patterns across hospitals and countries, the 4CE consortium is uniquely positioned to identify international differences in patient trajectories and hospital care that affect patient and population-level outcomes. To mitigate the intrinsic noisiness of EHR data, we performed extensive and iterative quality controls to address potential imprecision and incompleteness of datasets. Careful work with collaborators uncovered and addressed site-specific variations in data extraction and incomplete mapping of local EHR codes to desired data elements. Second, we used aggregate rather than patient-level data from each site for the current analysis in order to minimize privacy concerns and expedite the collaboration.

Aggregation enabled rapid sharing of data across sites and countries. As such, we could not assess the prediction performance of a composite risk score at patient level but instead report the predictive accuracy at the site level. To address this concern, we used comprehensive statistical tools to perform meta-analyses across sites. Third, the severity algorithm can accurately identify patients hospitalized with COVID-19 who develop severe disease, but it cannot presently identify the clinical trajectory that leads up to such a transition. Given the limitations of the severity algorithm that we previously validated, [8] we could not differentiate between patients who presented in a severe state and those who transitioned to severe disease over time. Our clinical experience and patient-level data from several institutions suggested that some patients were severe at admission while other patients transitioned to severe disease within a week of admission, sometimes suddenly. Thus, post-admission laboratory values might not capture the transition to severe disease.

### Unanswered questions and future research

We plan future analyses of patient-level data at each site to fully assess the temporal trend of individual and composite laboratory tests throughout the disease course. We have established a platform of harmonized data that is enabling our consortium to study questions related to coagulopathy, acute renal failure, pediatric symptomology, neurological sequelae, and many other areas of interest of our members. As the pandemic evolves, we are also examining how the data collection assumptions made in the first wave are faring when presentations of those with chronic COVID-19 disease is now present within our health systems. Our future work related to severity prediction will optimize risk scores based on laboratory values and other clinical features to capture temporal changes in patient population and clinical care. Additionally, we are evaluating the case mix changes for hospitals over time that may influence identification and type of severe COVID-19 disease.

### Meaning of the study: possible explanations and implications for clinicians and policymakers

We make several noteworthy observations. First, the key laboratory values emerging from our study represent the combination of acute inflammatory changes (CRP, LDH, procalcitonin) and underlying physiology (albumin). Both baseline health state and acuity of physiologic response to severe viral infection predict the risk of severe COVID-19. Second, the relatively low AUCs are likely due to the large variation within each site for a given laboratory value. When used in combination, prediction performance was dramatically improved. Third, the performance of the combined algorithm was relatively stable--and did not improve--over the first days of hospitalization. Stability over time implies that laboratory values at admission can be used to differentiate those patients likely to develop severe disease almost as well as models from subsequent days of hospitalization. As such, the current study examines the timing of laboratory tests in distinguishing patients who develop severe disease from those who do not.

Our systematic approach and extensive quality control allowed us to aggregate EHR data across sites and countries in a collaborative fashion at scale. The value of this work arises from leveraging these cross-country differences and similarities. While there may be dramatic differences in the outcomes of patients based on the complex interplay of comorbid conditions, natural history of the disease, and healthcare system dynamics, this fundamental similarity highlights a truism of disease: that there are clear and consistent patterns to the way humans react to disease. As we continue to build our collaborative and pursue future patient-level analyses, we will seek to uncover more subtle patterns and the role healthcare systems have in defining such perturbations.

## Data Availability

Only aggregate data was shared by sites for this study. All aggregate data in a de-identified fashion can be found and downloaded at www.covidclinical.net.

## Data Availability

Only aggregate data was shared by sites for this study. All aggregate data in a de-identified fashion can be found and downloaded at www.covidclinical.net. https://www.covidclinical.net

## SUMMARY BOX

Numerous studies have demonstrated that a significant proportion of hospitalized patients with COVID-19 develop severe disease with cardiorespiratory failure, requirements for mechanical ventilation and, sometimes, death. Numerous studies have developed prediction models using clinical data, including laboratory values, to predict patients at high risk of severe disease. However, most models have not been tested across hospital systems and multi-national cohorts to determine generalizability.

Across six countries and 46 health systems, we showed that average patient trajectories and their associations with severity risk were highly similar and consistent with previous models. Additionally, for most individual and combined laboratory value predictors (with the notable exception of D-dimer), initial average values were equivalent to those collected on subsequent days after admission.

## ETHICS STATEMENT

All study sites were responsible for and obtained ethics approval, as needed, from the appropriate ethics committee at their institution.

The lead author affirms that the manuscript is an honest, accurate, and transparent account of the study being reported; that no important aspects of the study have been omitted; and that any discrepancies from the study as originally planned have been explained.

The views expressed are those of the authors and do not necessarily represent the views or policy of the Department of Veterans Affairs or the United States Government.

## REFERENCES

1 Wu Z, McGoogan JM. Characteristics of and Important Lessons From the Coronavirus Disease 2019 (COVID-19) Outbreak in China: Summary of a Report of 72 314 Cases From the Chinese Center for Disease Control and Prevention. JAMA 2020;323:1239–42.

2 Wynants L, Van Calster B, Bonten MMJ, et al. Prediction models for diagnosis and prognosis of covid-19 infection: systematic review and critical appraisal. BMJ 2020;369:m1328.

3 Goyal P, Choi JJ, Pinheiro LC, et al. Clinical Characteristics of Covid-19 in New York City. N Engl J Med 2020;382:2372–4.

4 Fried MW, Crawford JM, Mospan AR, et al. Patient Characteristics and Outcomes of 11,721 Patients with COVID19 Hospitalized Across the United States. Clin Infect Dis Published Online First: 28 August 2020. doi:10.1093/cid/ciaa1268

5 Brat GA, Weber GM, Gehlenborg N, et al. International electronic health record-derived COVID-19 clinical course profiles: the 4CE consortium. NPJ Digit Med 2020;3:109.

6 Lippi G, Plebani M. Laboratory abnormalities in patients with COVID-2019 infection. Clinical Chemistry and Laboratory Medicine (CCLM). 2020;0. doi:10.1515/cclm-2020-0198

7 Chen X, Liu Z. Early prediction of mortality risk among severe COVID-19 patients using machine learning. Epidemiology. 2020. doi:10.1101/2020.04.13.20064329

8 Klann JG, Weber GM, Estiri H, et al. Validation of a Derived International Patient Severity Algorithm to Support COVID-19 Analytics from Electronic Health Record Data. Health Informatics. 2020;:e10020. doi:10.1101/2020.10.13.20201855

9 Phase2. 1SqlDataExtraction. Github https://github.com/covidclinical/Phase2.1SqlDataExtraction (accessed 2 Nov 2020).

10 DerSimonian R, Laird N. Meta-analysis in clinical trials. Control Clin Trials 1986;7:177–88.

11 COVID-19 Dashboard by the Center for Systems Science and Engineering (CSSE) at Johns Hopkins University (JHU). https://www.arcgis.com/apps/opsdashboard/index.html#/bda7594740fd40299423467b48e9ecf6 (accessed 7 Oct 2020).

12 Guan W-J, Ni Z-Y, Hu Y, et al. Clinical Characteristics of Coronavirus Disease 2019 in China. N Engl J Med Published Online First: 28 February 2020. doi:10.1056/NEJMoa2002032

