## Supplementary File for "International Comparisons of Harmonized Laboratory Value Trajectories to Predict Severe COVID-19: Leveraging the 4CE Collaborative Across 342 Hospitals and 6 Countries: A Retrospective Cohort Study" (from another version of this preprint: version 1, 2020-12-18)

I. **Further Methods**
   A. **AUC estimation for laboratory Tests**
II. **Efigures**
    A. **eFIGURE 1. Risk difference of severe disease between age groups 0-25, 26-49, 70-79, and 80+ compared to age group 50-69 stratified by country.**
    B. **eFIGURE 2: A) Pooled laboratory values in ever-severe and never-severe patients for selected laboratory tests, and B) patient-level AUC at each day after admission for those labs.**
    C. **eFIGURE 3: Site-level and country-level variation of laboratory tests.**
    D. **eFIGURE 4: Patient level AUC of laboratory tests stratified by regions.**
III. **Etables**
     A. **eTABLE1: Membership of 4CE.**
     B. **eTABLE2: Participating sites.**
     C. **eTABLE 3: Definition of severe COVID-19 based on the 4CE Severity Algorithm.**
     D. **eTABLE 4: Six comma-separated values (CSV) files and their descriptions.**
     E. **eTABLE 5: Pooled difference between admission laboratory measures in ever-severe and never-severe patients over all sites.**
I. **Further Methods**

**AUC estimation for laboratory tests**

We estimated patient-level AUC under distribution assumptions. The averaged AUC over sites on day $t$ was calculated as $\underline{AUC}_{jt}=\frac{\sum_{j=1}^{K} {\hat{w_{j} AUC}}_{jt}}{\sum_{j=1}^{K} w_{j}}$, where $\hat{{AUC}_{jt}}=\varphi(\frac{\mu_{jt1}-\mu_{jt0}}{\sqrt{\sigma_{jt1}^{2}+\sigma_{jt0}^{2}}})$was the estimated AUC at the $j$th site, $w_{j}$was the maximum number of patients at the $j$the site over 14 days, $\mu_{jt1}$, $\mu_{jt0}$, $\sigma_{jt1}$ and $\sigma_{jt0}$ were mean and standard deviation of lab measures among ever-severe patients and those among never-severe patients at the $j$the site for $j=1,...,K.$ We also calculated the standard error of $\underline{AUC}_{jt}$ via monte carlo simulation. Specifically, at the $j$th site on day $t$, we repeatedly generated the four measures: $\mu_{jt1}^{(d)}$, $\mu_{jt0}^{(d)}$, $\sigma_{jt1}^{2(d)}$ and $\sigma_{jt0}^{2(d)}$, for $d=1,...,1000$, where $\mu_{jty}^{(d)}$ was generated from a normal distribution $N(\mu_{ity}, \sigma_{jty}^{2}/n_{jty})$, and $\sigma_{jty}^{2(d)}$ was generated from a Chi-squared distribution ${\chi^{2}(n_{jty}-1)\sigma_{jty}^{2}/(n_{jty}-1)}_{{}}$ for $y=1,0$. We then calculated $\hat{{AUC}_{jt}^{(d)}}$ as the function of $\mu_{jt1}^{(d)}$, $\mu_{jt0}^{(d)}$, $\sigma_{jt1}^{2(d)}$ and $\sigma_{jt0}^{2(d)}$. The standard error of $\hat{{AUC}_{jt}}$ was then calculated as ${\hat{SE}_{jt}}=\sqrt{\frac{1}{1000}\sum_{d=1}^{1000} {\hat{(AUC}_{jt}^{(d)}-\frac{1}{1000}\sum_{d=1}^{1000} \hat{(AUC}_{jt}^{(d)})}^{2}}$. Finally, the averaged SE of AUC over sites on day $t$ was calculated as $\underline{SE}_{jt}=\frac{\sum_{j=1}^{K} w_{j}\hat{SE}_{jt}}{\sum_{j=1}^{K} w_{j}}$.

II. **eFIGURES**

**eFIGURE 1**. Risk difference of severe disease between age groups 0-25, 26-49, 70-79, and 80+ compared to age group 50-69 stratified by country. Risk is defined as the likelihood of being ever-severe.

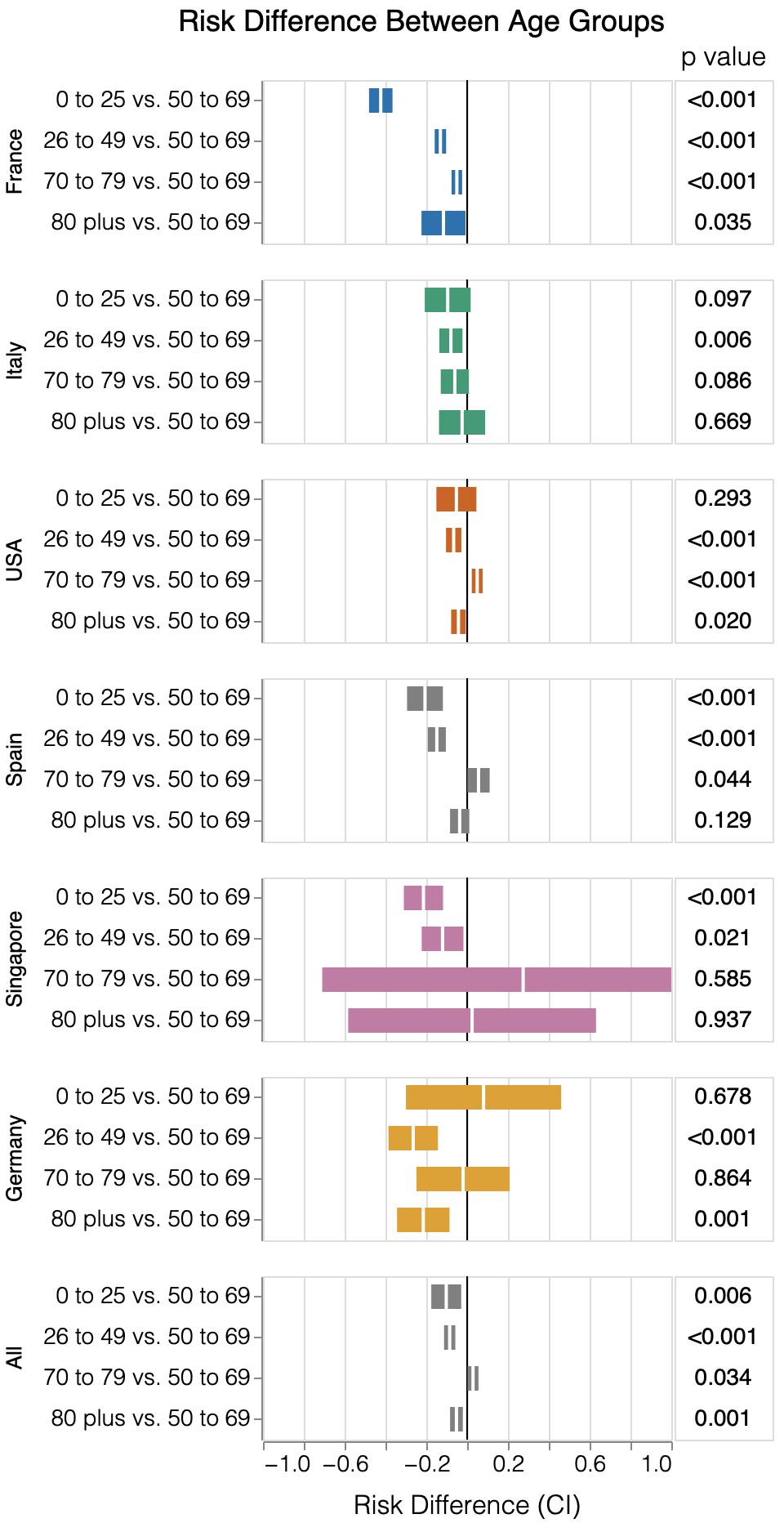

**eFIGURE 2**: A) Pooled laboratory values in ever-severe and never-severe patients for selected laboratory tests, and B) patient-level AUC at each day after admission for those labs. Inset shows AUC of laboratory value at admission and in-hospital to individually predict ever-severe as well as optimized thresholds.

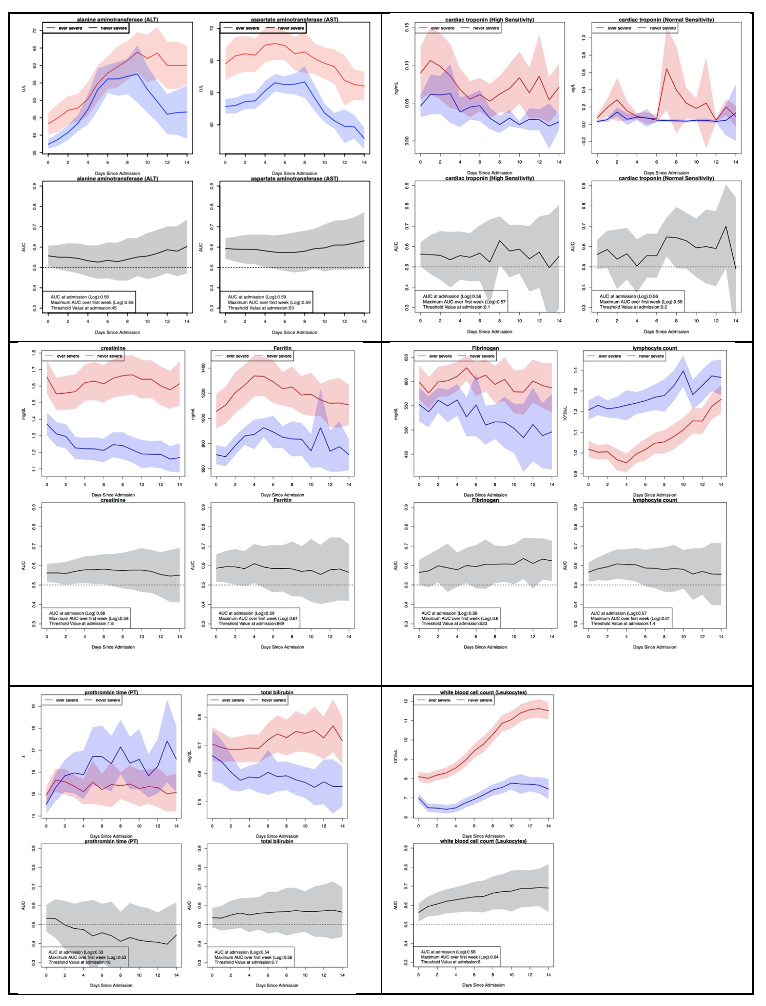

**eFIGURE 3**: Site-level and country-level variation of laboratory tests.

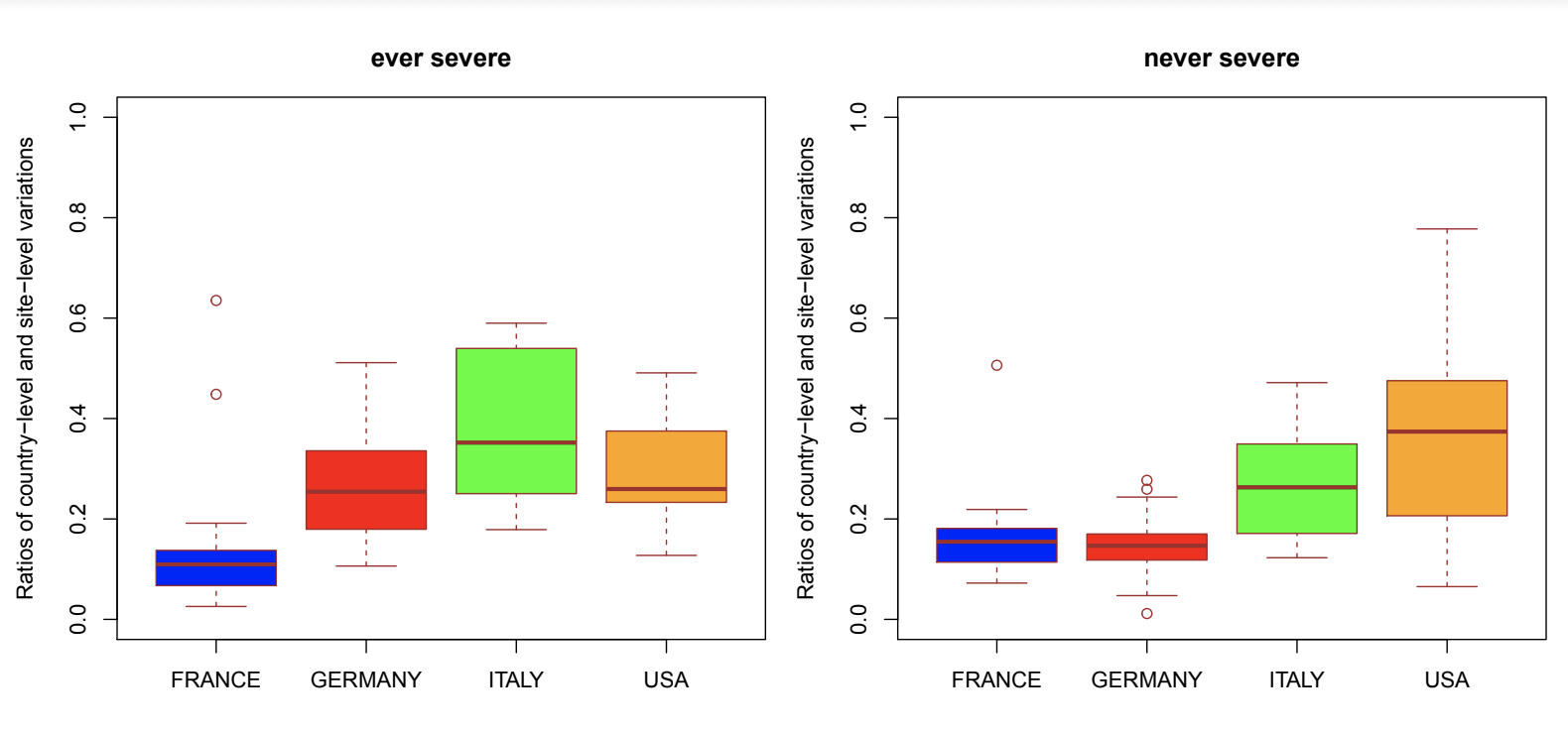

**eFIGURE 4**: Patient level AUC of laboratory tests stratified by regions.

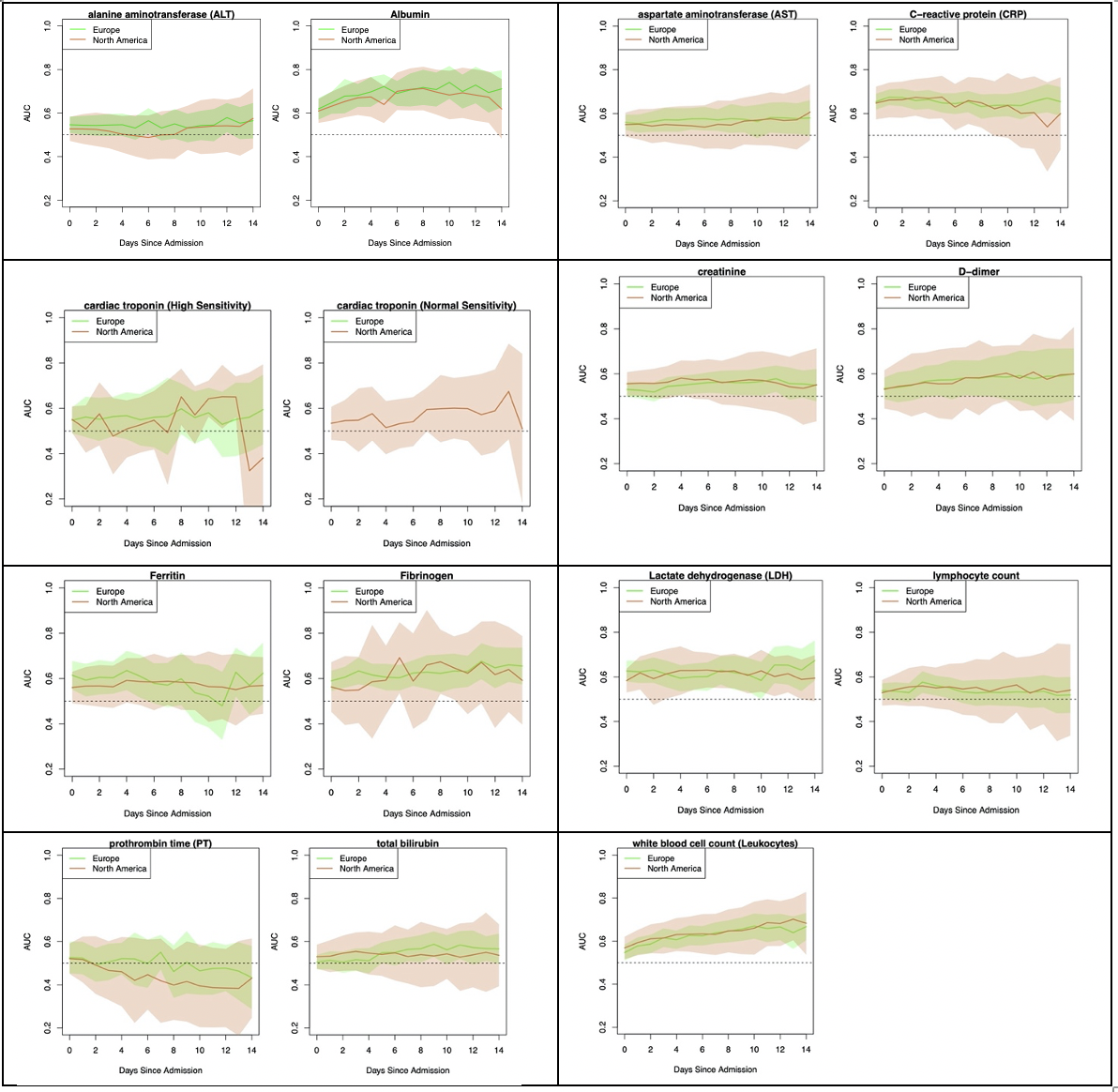

**IV. eTABLES**

**eTABLE1**: Membership of 4CE.

| First Name | **Middle Initial** | **Last Name** | **Degree(s)** | **Email Address** | **Primary Affiliation - Institution** | **Country** |
| --- | --- | --- | --- | --- | --- | --- |
| James | R | Aaron | MHA | | University of Kentucky | United States |
| Giuseppe |  | Agapito | PhD | | University Magna Graecia of Catanzaro, Italy | Italy |
| Adem |  | Albayrak |  | | Health Catalyst, INC. | United States |
| Mario |  | Alessiani | MD, FACS | | ASST Pavia, Lombardia Region Health System | Italy |
| Danilo | F | Amendola | MD | | Clinical Research Unit of Botucatu Medical School, São Paulo State University, Botucatu, Brazil | Brazil |
| Li | L.L.J | Anthony |  | | National Center for Infectious Diseases, Tan Tock Seng Hospital, Singapore | Singapore |
| Bruce | J | Aronow | PhD | | Cincinnati Children's Hospital Medical Center, University of Cincinnati | United States |
| Andrew |  | Atz | MD | | Medical University of South Carolina | United States |
| Paul |  | Avillach | MD, PhD | | Harvard Medical School | United States |
| James |  | Balshi |  | | St. Luke's University Health Network, Bethlehm PA | United States |
| Brett | K | Beaulieu-Jones | PhD | | Harvard Medical School | United States |
| Douglas | S | Bell |  | | Department of Medicine, David Geffen School of Medicine at UCLA | United States |
| Antonio |  | Bellasi |  | | UOC Ricerca, Innovazione e Brand reputation, ASST Papa Giovanni XXIII, Bergamo | Italy |
| Riccardo |  | Bellazzi | MS, PhD | | University of Pavia, Italy | Italy |
| Vincent |  | Benoit | PhD | | APHP Greater Paris University Hospital | France |
| Michele |  | Beraghi |  | | ASST Pavia | Italy |
| José Luis |  | Bernal Sobrino | MS | | Hospital Universitario 12 de Octubre, Madrid, Spain | Spain |
| Mélodie |  | Bernaux |  | | APHP Greater Paris University Hospital | France |
| Romain |  | Bey |  | | APHP Greater Paris University Hospital | France |
| Alvar |  | Blanco Martínez | MS | | Hospital Universitario 12 de Octubre, Madrid, Spain | Spain |
| Martin |  | Boeker |  | | Faculty of Medicine and Medical Center, University of Freiburg, Freiburg, Germany | Germany |
| Clara-Lea |  | Bonzel | MSc | | Harvard Medical School | United States |
| John |  | Booth | MSc | | Great Ormond Street Hospital for Children, UK | United Kingdom |
| Silvano |  | Bosari |  | | Scientific Direction, IRCCS Ca' Granda Ospedale Maggiore Policlinico di Milano | Italy |
| Florence | T | Bourgeois |  | | Harvard Medical School | United States |
| Robert | L | Bradford |  | | North Carolina Translational and Clinical Sciences (NC TraCS) Institute, UNC Chapel Hill | United States |
| Gabriel | A | Brat | MD | | Harvard Medical School | United States |
| Stéphane |  | Bréant |  | | APHP Greater Paris University Hospital | France |
| Mauro |  | Bucalo |  | | BIOMERIS (BIOMedical Research Informatics Solutions) | Italy |
| Anita |  | Burgun |  | | APHP Greater Paris University Hospital | France |
| Tianxi |  | Cai | PhD | | Harvard Medical School | United States |
| Mario |  | Cannataro | M.Sc. | | University Magna Graecia of Catanzaro, Italy | Italy |
| Aize |  | Cao |  | | Tennessee Valley Healthcare System Veterans Affairs Medical Center | United States |
| Aldo |  | Carmona |  | | Department of Anesthesia, St. Luke's University Health Network, Bethlehem, PA | United States |
| Charlotte |  | Caucheteux |  |  | Université Paris-Saclay, Inria, CEA | France |
| Julien |  | Champ |  | | INRIA Sophia-Antipolis – ZENITH team, LIRMM, Montpellier, France | France |
| Jin |  | Chen | PhD | | University of Kentucky | United States |
| Luca |  | Chiovato |  | | Unit of Internal Medicine and Endocrinology, Istituti Clinici Scientifici Maugeri SpA SB IRCCS, Pavia, Italy and Department of Internal Medicine and Therapeutics, University of Pavia, Pavia, Italy. | Italy |
| James | J | Cimino |  | | UAB Informatics Institute | United States |
| Tiago | K | Colicchio |  | | UAB Informatics Institute | United States |
| Sylvie |  | Cormont |  | | APHP Greater Paris University Hospital | France |
| Sébastien |  | Cossin |  | | Bordeaux University Hospital / ERIAS - Inserm U1219 BPH | France |
| Jean | B | Craig | PhD | | Medical University of South Carolina | United States |
| Juan Luis |  | Cruz Bermúdez | PhD | | Hospital Universitario 12 de Octubre, Madrid, Spain | Spain |
| Jaime |  | Cruz Rojo | MD | | Hospital Universitario 12 de Octubre, Madrid, Spain | Spain |
| Arianna |  | Dagliati | MS, PhD | | University of Pavia, Italy | Italy |
| Mohamad |  | Daniar | MSIS | | Boston Children's Hospital | United States |
| Christel |  | Daniel |  | | APHP Greater Paris University Hospital, INSERM | France |
| Anahita |  | Davoudi | PhD | | University of Pennsylvania Perelman School of Medicine | United States |
| Batsal |  | Devkota |  | | Department of Biomedical and Health Informatics, Children's Hospital of Philadelphia | United States |
| Julien |  | Dubiel |  |  | APHP Greater Paris University Hospital | France |
| Scott | L | DuVall |  | | VA Informatics and Computing Infrastructure, VA Salt Lake City Health Care System | United States |
| Loic |  | Esteve |  |  | SED/SIERRA, Inria Centre de Paris | France |
| Shirley |  | Fan |  | | University of Michigan | United States |
| Robert | W | Follett |  | | Department of Medicine, David Geffen School of Medicine at UCLA | United States |
| Paula | S. A | Gaiolla |  | | Internal Medicine Department of Botucatu Medical School, São Paulo State University, Botucatu, Brazil | Brazil |
| Thomas |  | Ganslandt | MD | | University Medicine Mannheim, Heidelberg University | Germany |
| Noelia |  | García Barrio | MS | | Hospital Universitario 12 de Octubre, Madrid, Spain | Spain |
| Nils |  | Gehlenborg |  | | Harvard Medical School | United States |
| Alon |  | Geva | MD, MPH | | Boston Children's Hospital, Boston, MA | United States |
| Tobias |  | Gradinger | MD, BSc | | University Medicine Mannheim, Heidelberg University | Germany |
| Alexandre |  | Gramfort |  | | Université Paris-Saclay, Inria, CEA | France |
| Romain |  | Griffier |  | | Bordeaux University Hospital | France |
| Nicolas |  | Griffon |  | | APHP Greater Paris University Hospital, INSERM | France |
| Olivier |  | Grisel |  |  | Université Paris-Saclay, Inria, CEA | France |
| Alba |  | Gutiérrez-Sacristán | PhD | | Harvard Medical School | United States |
| David | A | Hanauer | MD, MS | | University of of Michigan Medical School, Ann Arbor, Michigan, USA | United States |
| Christian |  | Haverkamp |  |  | Institute of Digitalization in Medicine, Faculty of Medicine and Medical Center, University of Freiburg, Germany | Germany |
| Darren | W | Henderson |  | | University of Kentucky | United States |
| Martin |  | Hilka |  |  | APHP Greater Paris University Hospital | France |
| John | H | Holmes | MS, PhD | | University of Pennsylvania Perelman School of Medicine (inst), Philadelphia, Pennsylvania, United States of America | United States |
| Chuan |  | Hong | PhD | | Harvard Medical School | United States |
| Petar |  | Horki |  | | Institute of Medical Biometry and Statistics, Medical Center, University of Freiburg | Germany |
| Meghan | R | Hutch | BS | | Northwestern University | United States |
| Richard | W | Issitt | DClinP | | Great Ormond Street Hospital for Children, UK | United Kingdom |
| Anne Sophie |  | Jannot |  | | Department of Biomedical Informatics, HEGP,  APHP Greater Paris University Hospital | France |
| Vianney |  | Jouhet | MD,PhD | | Bordeaux University Hospital | France |
| Mark | S | Keller |  | | Harvard Medical School | United States |
| Katie |  | Kirchoff | MSHI | | Medical University of South Carolina | United States |
| Jeffrey | G | Klann | MEng, PhD | | Massachusetts General Hospital | United States |
| Isaac | S | Kohane | MD, PhD | | Harvard Medical School | United States |
| Ian | D | Krantz |  | | Department of Pediatrics, Division of Human Genetcs, The Children's Hospital of Philadelphia and the Perelman School of Medicine at the University of Pennsylvania | United States |
| Detlef |  | Kraska | Dr. | | University Hospital Erlangen | Germany |
| Ashok | K | Krishnamurthy | PhD | | University of North Carolina, Chapel Hill | United States |
| Sehi |  | L'Yi | PhD | | Harvard Medical School | United States |
| Trang | T | Le | PhD | | University of Pennsylvania Perelman School of Medicine | United States |
| Judith |  | Leblanc |  | | APHP Greater Paris University Hospital | France |
| Guillaume |  | Lemaitre |  |  | Université Paris-Saclay, Inria, CEA | France |
| Leslie |  | Lenert | MD, MS | | Medical University of South Carolina | United States |
| Damien |  | Leprovost |  |  | Clevy.io | France |
| Molei |  | Liu | PhD | | Harvard T.H. Chan School of Public Health | United States |
| Ne Hooi Will |  | Loh | MBBS | | National University Health System, Singapore | Singapore |
| Yuan |  | Luo | PhD | | Northwestern University | United States |
| Kristine | E | Lynch |  | | VA Informatics and Computing Infrastructure, VA Salt Lake City Health Care System | United States |
| Sadiqa |  | Mahmood |  | | Health Catalyst, INC. | United States |
| Sarah |  | Maidlow |  | | University of Michigan | United States |
| Alberto |  | Malovini | PhD | | Istituti Clinici Scientifici Maugeri SpA SB IRCCS, Pavia, Italy. | Italy |
| Kenneth | D | Mandl | MD, MPH | | Boston Children's Hospital | United States |
| Chengsheng |  | Mao | PhD | | Northwestern University | United States |
| Patricia |  | Martel |  | | APHP Greater Paris University Hospital | France |
| Aaron | J | Masino | PhD | | Children's Hospital of Philadelphia | United States |
| Michael | E | Matheny |  | | Tennessee Valley Healthcare System Veterans Affairs Medical Center | United States |
| Thomas |  | Maulhardt |  | | University Medical Center Freiburg, Germany | Germany |
| Maria |  | Mazzitelli | PhD | | University Magna Graecia of Catanzaro, Italy | Italy |
| Michael | T | McDuffie |  | | Harvard Medical School | United States |
| Arthur |  | Mensch |  |  | ENS, PSL University | France |
| Marianna |  | Milano | PhD | | University Magna Graecia of Catanzaro, Italy | Italy |
| Marcos | F | Minicucci |  |  | Internal Medicine Department of Botucatu Medical School, São Paulo State University, Botucatu, Brazil | Brazil |
| Bertrand |  | Moal | PhD | | Bordeaux University Hospital | France |
| Jason | H | Moore | PhD | | University of Pennsylvania Perelman School of Medicine | United States |
| Jeffrey | S | Morris |  | | University of Pennsylvania Perelman School of Medicine | United States |
| Michele |  | Morris | BA | | University of Pittsburgh | United States |
| Karyn | L | Moshal |  | | Great Ormond Street Hospital for Children, UK | United Kingdom |
| Sajad |  | Mousavi | PhD | | Harvard Medical School | United States |
| Danielle | L | Mowery | PhD | | University of Pennsylvania Perelman School of Medicine | United States |
| Douglas | A | Murad |  | | David Geffen School of Medicine at UCLA | United States |
| Shawn | N | Murphy | MD, PhD |, googledoc is | Massachusetts General Hospital | United States |
| Thomas | P | Naughton | BA | | Harvard Medical School | United States |
| Antoine |  | Neuraz | MD, PhD | | Hôpital Necker-Enfants Malade, Assistance Publique Hôpitaux de Paris (APHP), University of Paris | France |
| Kee Yuan |  | Ngiam | MBBS, MRCS, MMed, FRCS | | National Univerisity Health Systems Singapore | Singapore |
| Jihad |  | Obeid | MD, FAMIA | | Medical University of South Carolina | United States |
| Marina | P | Okoshi | PhD | | Internal Medicine Department of Botucatu Medical School, São Paulo State University, Botucatu, Brazil | Brazil |
| Karen | L | Olson | PhD | | Boston Children's Hospital and Harvard Medical School | United States |
| Gilbert | S. | Omenn | MD, PhD | | University of Michigan | United States |
| Nina |  | Orlova |  |  | APHP Greater Paris University Hospital | France |
| Brian | D | Ostasiewski |  | | CTSI, WFBMI, Wake Forest School of Medicine | United States |
| Nicolas |  | Paris |  | | APHP Greater Paris University Hospital | France |
| Lav | P | Patel | MS | | University Of Kansas Medical Center | United States |
| Miguel |  | Pedrera Jimenez | MS | | Hospital Universitario 12 de Octubre, Madrid, Spain | Spain |
| Nathan | P | Palmer | PhD | | Harvard Medical School | United States |
| Danielle |  | Pillion | MS | | Harvard Medical School | United States |
| Hans | U | Prokosch |  | | University of Erlangen-Nürnberg | Germany |
| Robson | A | Prudente | PhD | | Clinical Research Unit São Paulo State University, Brazil | Brazil |
| Víctor |  | Quirós González | MS | | Hospital Universitario 12 de Octubre, Madrid, Spain | Spain |
| Rachel | B | Ramoni |  | | Office of Research and Development, Department of Veterans Affairs | United States |
| Maryna |  | Raskin |  | | Health Catalyst, INC. | United States |
| Siegbert |  | Rieg |  | | Division of Infectious Diseases, Department of Medicine II, Medical Center – University of Freiburg, Faculty of Medicine | Germany |
| Gustavo |  | Roig Domínguez | MS | | Hospital Universitario 12 de Octubre, Madrid, Spain | Spain |
| Elisa |  | Salamanca |  | | APHP Greater Paris University Hospital | France |
| Malarkodi | J | Samayamuthu | MD | | University of Pittsburgh | United States |
| Arnaud |  | Sandrin |  | | APHP Greater Paris University Hospital | France |
| Maria |  | Savino | MS | | Management Engineer, Direction | Italy |
| Emily | R | Schriver | MS | | University of Pennsylvania Health System | United States |
| Juergen |  | Schuettler |  | | Department of Anesthesiology, University Hospital Erlangen, FAU Erlangen-Nürnberg, Germany | Germany |
| Luigia |  | Scudeller |  | | Scientific Direction, IRCCS Ca' Granda Ospedale Maggiore Policlinico di Milano | Italy |
| Neil | J | Sebire | MD, FRCPath | | Great Ormond Street Hospital for Children NIHR BRC, UK | United Kingdom |
| Pablo |  | Serrano Balazote | MD,MS | | Hospital Universitario 12 de Octubre, Madrid, Spain | Spain |
| Patricia |  | Serre |  |  | APHP Greater Paris University Hospital | France |
| Arnaud |  | Serret-Larmande | MD | |  | France |
| Domenick |  | Silvio |  | | University of Michigan, MICHR | United States |
| Piotr |  | Sliz |  | | Boston Children's Hospital | United States |
| Jiyeon |  | Son | MD | | University of Pittsburgh Medical Center | United States |
| Charles |  | Sonday |  | | Critical Care Medicine, Department of Medicine, St. Luke's University Health Network, Bethlehm PA | United States |
| Andrew | M | South | MD, MS | | Brenner Children's Hospital, Wake Forest School of Medicine | United States |
| Anastasia |  | Spiridou | PhD, MSc | | Great Ormond Street Hospital for Children, UK | United Kingdom |
| Amelia | LM | Tan | BSc, PhD | | Harvard Medical School | United States |
| Bryce | W.Q. | Tan | MBBS | | National University Hospital, Singapore | Singapore |
| Byorn | W.L. | Tan | MBBS | | National University Hospital, Singapore | Singapore |
| Suzana | E | Tanni |  | | Internal Medicine Department of Botucatu Medical School, São Paulo State University, Botucatu, Brazil | Brazil |
| Deanne | M | Taylor | PhD | | The Children's Hospital of Philadelphia and the University of Pennsylvania Perelman Medical School | United States |
| Ana | I | Terriza Torres | MS | | Hospital Universitario 12 de Octubre, Madrid, Spain | Spain |
| Valentina |  | Tibollo |  | | Laboratory of Informatics and Systems Engineering for Clinical Research, Istituti Clinici Scientifici Maugeri SpA SB IRCCS, Pavia, Italy. | Italy |
| Patric |  | Tippmann | MSc | | Institute of Medical Biometry and Statistics, Medical Center, University of Freiburg | Germany |
| Carlo |  | Torti | PhD | | University Magna Graecia of Catanzaro, Italy | Italy |
| Enrico | M | Trecarichi | PhD | | University Magna Graecia of Catanzaro, Italy | Italy |
| Andrew | K | Vallejos |  | | Clinical & Translational Science Institute, Medical College of Wisconsin | United States |
| Gael |  | Varoquaux |  |  | Université Paris-Saclay, Inria, CEA, Montréal Neurological Institute, McGill University | France |
| Jill-Jênn |  | Vie |  |  | SequeL, Inria Lille | France |
| Shyam |  | Visweswaran | MD, PhD | | University of Pittsburgh | United States |
| Michele |  | Vitacca | MD, PhD | | ICS S. Maugeri IRCCS Pavia Italy | ITALY |
| Kavishwar | B | Wagholikar | MBBS, PhD | | Department of Medicine, Massachusetts General Hospital, Boston, USA | United States |
| Lemuel | R | Waitman |  | | University of Missouri, Columbia. MO | United States |
| Demian |  | Wassermann |  |  | Université Paris-Saclay, Inria, CEA | France |
| Griffin | M | Weber |  | | Harvard Medical School | United States |
| Yuan |  | William |  | | Harvard Medical School | United States |
| Zongqi |  | Xia | MD, PhD | | University of Pittsburgh | United States |
| Nadir |  | Yehya | MD, MSCE | | Children's Hospial of Philadelphia and University of Pennsylvania | United States |
| Alberto |  | Zambelli |  | | ASST Papa Giovanni XXIII, Bergamo | Italy |
| Harrison | G | Zhang |  | | Harvard Medical School | United States |
| Chiara |  | Zucco | PhD | | University Magna Graecia of Catanzaro, Italy | Italy |
| James | B | Norman |  | | Harvard Medical School | United States |
| Anupama |  | Maram | MS | | Harvard Medical School | United States |
| Nicholas | W | Brown | MEng | | Harvard Medical School | United States |
| Kenneth | M | Huling | HS | | Harvard Medical School | United States |
| Emily | R | Pfaff | PhD | | UNC Chapel Hill | United States |
| Lorenzo |  | Chiudinelli | PhD | | ASST Papa Giovanni XXIII, Bergamo | Italy |
| Yi-Ju |  | Tseng | PhD | | Chang Gung University | Taiwan |

**eTABLE2**: Participating sites.

| **Healthcare System** | **Acronym** | **City** | **Country** | **Patient Population** | **Hospitals** | **Beds** | **Inpatient discharges/year** | **Obfuscation small count mask threshold (e.g., <10)** |
| --- | --- | --- | --- | --- | --- | --- | --- | --- |
| Assistance Publique - Hôpitaux de Paris | APHP | Paris | France | Adult & Pediatric | 39 | 20098 | 1375538 | <3 |
| Bordeaux University Hospital | FRBDX | Bordeaux | France | Adult & Pediatric | 3 | 2676 | 130033 | <3 |
| Erlangen University Hospital | UKER | Erlangen | Germany | Adult & Pediatric | 1 | 1400 | 65000 | <5 |
| Medical Center, University of Freiburg | UKFR | Freiburg | Germany | Adult & Pediatric | 1 | 1660 | 71500 | <4 |
| University Medicine Mannheim | UMM | Mannheim | Germany | Adult & Pediatric | 1 | 1352 | 50748 | <3 |
| ICSM Pavia Hospital | ICSM1 | Pavia | Italy | Adult | 1 | 426 | 8616 | none |
| ICSM Lumezzane/Brescia Hospitals | ICSM5 | Lumezzane/Brescia | Italy | Adult | 1 | 149 | 1296 | none |
| ICSM Milano Hospital | ICSM20 | Milan | Italy | Adult | 1 | 200 | 2432 | none |
| Policlinico di Milano | POLIMI | Milan | Italy | Adult & Pediatric | 1 | 900 | 40000 | <5 |
| ASST Papa Giovanni XXIII Bergamo | HPG23 | Bergamo | Italy | Adult & Pediatric | 1 | 1080 | 45000 | <=5 |
| National University Hospital | NUH | Singapore | Singapore | Adult & Pediatric | 1 | 1556 | 100977 | none |
| Beth Israel Deaconess Medical Center | BIDMC | Boston, MA | USA | Adult | 1 | 673 | 40752 | <10 |
| University of Kansas Medical Center | KUMC | Kansas City, KS | USA | Adult & Pediatric | 1 | 794 | 54659 | <=10 |
| Mass General Brigham (Partners Healthcare) | MGB | Boston, MA | USA | Adult & Pediatric | 10 | 3418 | 163521 | <10 |
| Medical University of South Carolina | MUSC | Charleston, SC | USA | Adult & Pediatric | 8 | 1600 | 55664 | <15 |
| University of Pennsylvania | UPenn | Philadelphia, PA | USA | Adult | 5 | 2469 | 118188 | <10 |
| University of California, LA | UCLA | Los Angeles, CA | USA | Adult & Pediatric | 2 | 786 | 40526 | <10 |
| University of Michigan | UMICH | Ann Arbor, MI | USA | Adult & Pediatric | 3 | 1000 | 49008 | none |
| University of North Carolina at Chapel Hill | UNC | Chapel Hill, NC | USA | Adult & Pediatric | 11 | 3095 | 52000 | <10 |
| UT Southwestern Medical Center | UTSW | Dallas, TX | USA | Adult | 1 | 608 | 26905 | <6 |
| Northwestern University | NWU | Chicago, IL | USA | Adult & Pediatric | 10 | 2234 | 103279 |  |
| University of Pittsburgh / UPMC | UPITT | Pittsburgh, PA | USA | Adult & Pediatric | 39 | 8085 | 369300 | none |
| Medical College of Wisconsin | MCWCTSI | Milwaukee, WI | USA | Adult | 3 | 876 | 44655 | <11 |
| Hospital Universitario 12 de Octubre | H12O | Madrid | Spain | Adult & Pediatric | 1 | 1256 | 45035 | <10 |
| Wake Forest School of Medicine & Brenner Children's Hospital | WFSM | Winston Salem, NC | USA | Adult & Pediatric | 5 | 1536 | 67541 | <10 |
| University Magna Graecia of Catanzaro & Mater Domini University Hospital, Catanzaro | UNICZ | Catanzaro | Italy | Adult | 1 | 210 | 11987 | none |
| St. Luke's University Health Network | SLHN | Bethlehem, PA | USA | Adult & Pediatric | 12 | 1700 | 75000 |  |
| University of Kentucky | UKY | Lexington, KY | USA | Adult & Pediatric | 3 | 881 | 45714 | none |
| Riverside Health System | RIVHS | Newport News, VA | USA | Adult & Pediatric | 5 | 670 | 30000 |  |
| ASST della provincia di Pavia | ASSTPAVIA | Pavia | Italy | Adult & Pediatric | 7 | 958 | 29103 | none |
| VISN1 - VA New England Health Care System | VA1 | Bedford, MA | USA | Adult | 11 |  |  | <10 |
| VISN2 - VA Healthcare New York/New Jersey | VA2 | Bronx, NY | USA | Adult | 14 |  |  | <10 |
| VISN4 - VISN4 network (PA, DE, NJ, OH) | VA4 | Pittsburgh, PA | USA | Adult | 11 |  |  | <10 |
| VISN5 - VA Capitol Healthcare Network | VA5 | Linthicum, MD | USA | Adult | 5 |  |  | <10 |
| VISN6 - VA Mid-Atlantic Health Care Network | VA6 | Durgham, NC | USA | Adult | 8 |  |  | <10 |
| VISN7 - VA Southeast Network | VA7 | Duluth, GA | USA | Adult | 10 |  |  | <10 |
| VISN8 - VA Sunshine Healthcare Network | VA8 | Saint Petersburg, FL | USA | Adult | 10 |  |  | <10 |
| VISN9 - VA MidSouth Healthcare Network | VA9 | Nashville, TN | USA | Adult | 9 |  |  | <10 |
| VISN10 - VA Healthcare System VISN10 | VA10 | Cincinnati, OH | USA | Adult | 15 |  |  | <10 |
| VISN12 - VA Great Lakes Healthcare System | VA12 | Hines, IL | USA | Adult | 6 |  |  | <10 |
| VISN15 - VA Heartland Network | VA15 | Kansas City, MO | USA | Adult | 9 |  |  | <10 |
| VISN16 - South Central VA Healthcare Network | VA16 | Ridgeland, MS | USA | Adult | 11 |  |  | <10 |
| VISN17 - VA Heart of Texas Healthcare System | VA17 | Arlington, TX | USA | Adult | 7 |  |  | <10 |
| VISN19 - VA Rocky Mountain Network | VA19 | Glendale, CO | USA | Adult | 6 |  |  | <10 |
| VISN20 - VA Northwest Health Network | VA20 | Vancouver, WA | USA | Adult | 8 |  |  | <10 |
| VISN21 - VA Sierra Pacific Network | VA21 | Mare Island, CA | USA | Adult | 9 |  |  | <10 |
| VISN22 - VA Desert Pacific Healthcare Network | VA22 | Long Beach, CA | USA | Adult | 5 |  |  | <10 |
| VISN23 - VA Midwest Healthcare Network | VA23 | Minneapolis, MN | USA | Adult | 9 |  |  | <10 |

**eTABLE 3**: Definition of severe COVID-19 based on the 4CE Severity Algorithm. Please refer to Klann et al. for further information.

| **EHR Code Category** | **Clinical Events** |
| --- | --- |
| Diagnoses | Acute respiratory distress syndrome, ventilator-associated pneumonia |
| Procedures | Insertion of endotracheal tube; invasive mechanical ventilation |
| Laboratory results | PaCO2, PaO2 |
| Anesthetic Medications | General anesthetics: ketamine, propofol  Benzodiazepine derivatives: midazolam  Muscle relaxants: cisatracurium, rocuronium, vecuronium  Other hypnotics and sedatives: dexmedetomidine |
| Vasopressor Medications | Adrenergic and dopaminergic agents: Dopamine, Epinephrine, Norepinephrine, Phenylephrine, Milrinone, Dobutamine  Other cardiac stimulants: Angiotensin II  Other respiratory system products:  phosphodiesterase inhibitors, Epoprostenol, Nitric Oxide  Vasopressin and analogs: Vasopressin |

**eTABLE 4**: Six comma-separated values (CSV) files and their descriptions. Sites optionally obfuscated the values in any of these files by replacing small counts with “-99.” Sites indicated missing data or data that they were unable to obtain (e.g., race and ethnicity not collected at some sites) with “-999.”

| **Data File** | **Description** |
| --- | --- |
| DailyCounts.csv | Contained one row per calendar date. Each row included the date, the cumulative number of patients admitted by this date, the cumulative number of patients who had developed severe disease by this date, the cumulative number of patients who had died by this date, the number of patients in the hospital on this date (i.e., excluding patients who have been discharged or have died), and the number of patients who became severe and are in the hospital on this date. |
| ClinicalCourse.csv | Contained one row per day since admission. For example, the first row contained aggregate data about patients on their admission date, and the second row contained aggregate data on the day following their admission. Each row indicated the number of total patients in the cohort and the number of severe patients in the hospital on that day. Patients who were discharged and then readmitted were counted for each day they were an inpatient. |
| Demographics.csv | Contained breakdown counts for both the total cohort and the severe patients by sex, age group (0-2, 3-5, 6-11, 12-17, 18-25, 26-49, 50-69, 70-79, and 80+ years old), and race and ethnicity category (American Indian or Alaska Native, Asian, Black or African American, Hispanic or Latino, Native Hawaiian or Other Pacific Islander, White) [https://grants.nih.gov/grants/guide/notice-files/not-od-15-089.html]. Reporting of the race categories was mostly restricted to the United States sites. |
| Labs.csv | Contained the daily trajectories of select laboratory tests. Each row corresponded to a laboratory test (identified using a LOINC code) and the number of days since admission. The values in each row were the number of patients who had a test result on that day, the mean and standard deviation of the test results, and the mean and standard deviation of the natural logarithm of the test results plus 0.5 (to avoid log zero). These statistics were provided for both the total cohort and the severe patients. If a patient had multiple results for the same test on the same day, then those were replaced by the mean of that day’s test results before averaging across all patients. Log mean values are more meaningful summaries for tests such as D-dimer with highly skewed distributions. |
| Diagnoses.csv | Contained one row per ICD9 or ICD10 diagnosis code, truncated to just their first 3 characters. This was a basic approach to rolling up diagnoses into broader categories and reducing the effect of coding differences between sites. For each truncated diagnosis code, the number of total and severe patients before and since admission with the code were listed. Before admission meant day -365 to day -15 relative to the index admission date. The two weeks immediately prior to admission were ignored to ensure that these diagnoses occurred before patients had symptoms concerning for COVID-19. Since admission meant day 0, the index date, until the day when the data were extracted. |
| Medications.csv | The sixth file, Medications.csv, had one row per each of 12 medication classes. For each medication class, the number of total and severe patients before and since admission for whom the medication was ordered were listed. Similar to diagnoses, before meant day -365 to -15, and since meant day 0+ relative to the index date. |

**eTABLE5**: Pooled difference between admission laboratory measures in ever-severe and never-severe patients over all sites.

|  | | **Difference of Laboratory Measures** | | | |
| --- | --- | --- | --- | --- | --- |
| **Scale** | **Labs** | **EST** | **SE** | **CI_95L** | **CI_95U** |
| original | alanine aminotransferase (ALT) | 5.17 | 1.24 | 2.73 | 7.61 |
|  | albumin | -0.21 | 0.02 | -0.24 | -0.17 |
|  | aspartate aminotransferase (AST) | 12.11 | 1.86 | 8.47 | 15.76 |
|  | C-reactive protein (CRP) (Normal Sensitivity) | 48.72 | 3.67 | 41.53 | 55.92 |
|  | cardiac troponin (High Sensitivity) | 0.03 | 0.03 | -0.02 | 0.09 |
|  | cardiac troponin (Normal Sensitivity) | 0.01 | 0.01 | 0.00 | 0.03 |
|  | creatinine | 0.24 | 0.04 | 0.17 | 0.32 |
|  | D-dimer | 171.14 | 39.12 | 94.47 | 247.80 |
|  | Ferritin | 299.10 | 53.50 | 194.24 | 403.95 |
|  | Fibrinogen | 46.75 | 7.98 | 31.12 | 62.38 |
|  | lactate dehydrogenase (LDH) | 116.86 | 13.87 | 89.69 | 144.04 |
|  | lymphocyte count | -0.17 | 0.02 | -0.21 | -0.13 |
|  | neutrophil count | 1.05 | 0.07 | 0.92 | 1.18 |
|  | procalcitonin | 0.51 | 0.09 | 0.34 | 0.68 |
|  | prothrombin time (PT) | 0.45 | 0.15 | 0.16 | 0.74 |
|  | total bilirubin | 0.05 | 0.01 | 0.03 | 0.07 |
|  | white blood cell count (Leukocytes) | 1.08 | 0.10 | 0.88 | 1.27 |
| log | alanine aminotransferase (ALT) | 0.13 | 0.02 | 0.09 | 0.17 |
|  | albumin | -0.05 | 0.01 | -0.06 | -0.04 |
|  | aspartate aminotransferase (AST) | 0.21 | 0.02 | 0.17 | 0.25 |
|  | C-reactive protein (CRP) (Normal Sensitivity) | 0.70 | 0.04 | 0.61 | 0.79 |
|  | cardiac troponin (High Sensitivity) | 0.05 | 0.01 | 0.02 | 0.08 |
|  | cardiac troponin (Normal Sensitivity) | 0.04 | 0.01 | 0.01 | 0.06 |
|  | creatinine | 0.10 | 0.01 | 0.08 | 0.12 |
|  | D-dimer | 0.26 | 0.06 | 0.14 | 0.39 |
|  | Ferritin | 0.37 | 0.05 | 0.27 | 0.47 |
|  | Fibrinogen | 0.07 | 0.02 | 0.04 | 0.10 |
|  | lactate dehydrogenase (LDH) | 0.27 | 0.03 | 0.21 | 0.32 |
|  | lymphocyte count | -0.10 | 0.01 | -0.12 | -0.08 |
|  | neutrophil count | 0.17 | 0.01 | 0.14 | 0.19 |
|  | procalcitonin | 0.29 | 0.03 | 0.24 | 0.34 |
|  | prothrombin time (PT) | 0.03 | 0.01 | 0.02 | 0.04 |
|  | total bilirubin | 0.04 | 0.00 | 0.03 | 0.05 |
|  | white blood cell count (Leukocytes) | 0.12 | 0.01 | 0.10 | 0.14 |

**eTABLE6:** IRB Approval Across Data-Contributing Sites

| **Institutional Review Board** | **Determination** |
| --- | --- |
| Assistance Publique - Hôpitaux de Paris | Approved |
| Bordeaux University Hospital | Exempt |
| Erlangen University Hospital | Approved |
| Medical Center, University of Freiburg | Exempt |
| University Medicine Mannheim | Approved |
| Instituti Clinici Scientifici Maugeri (ICSM) | Waived |
| Policlinico di Milano | Approved |
| ASST Papa Giovanni XXIII Bergamo | Approved |
| National University Hospital | Waived |
| Beth Israel Deaconess Medical Center | Exempt |
| University of Kansas Medical Center | Approved |
| Mass General Brigham (Partners Healthcare) | Exempt |
| Medical University of South Carolina | Waived |
| University of Pennsylvania | Exempt |
| University of California, LA | Exempt |
| University of Michigan | Exempt |
| UT Southwestern Medical Center | Approved |
| Northwestern University | Approved |
| University of Pittsburgh / UPMC | Approved |
| Hospital Universitario 12 de Octubre | Approved |
| Wake Forest School of Medicine & Brenner Children's Hospital | Approved |
| University Magna Graecia of Catanzaro & Mater Domini University Hospital | Approved |
| St. Luke's University Health Network | Waived |
| VA New England Health Care System | Approved |
| VA Healthcare New York/New Jersey | Approved |
| VISN4 network (PA, DE, NJ, OH) | Approved |
| VA Capitol Healthcare Network | Approved |
| VA Mid-Atlantic Health Care Network | Approved |
| VA Southeast Network | Approved |
| VA Sunshine Healthcare Network | Approved |
| VA MidSouth Healthcare Network | Approved |
| VA Healthcare System VISN10 | Approved |
| VA Great Lakes Healthcare System | Approved |
| VA Heartland Network | Approved |
| South Central VA Healthcare Network | Approved |
| VA Heart of Texas Healthcare System | Approved |
| VA Rocky Mountain Network | Approved |
| VA Northwest Health Network | Approved |
| VA Sierra Pacific Network | Approved |
| VA Desert Pacific Healthcare Network | Approved |
| VA Midwest Healthcare Network | Approved |
